# Conversions between noise exposure metrics 24-hour Leq, Ldn, and Lden: the impact of diurnal local bus traffic patterns on population annoyance in the United States

**DOI:** 10.1101/2023.10.25.23297557

**Authors:** Edmund Seto, Ching-Hsuan Huang

## Abstract

Noise during evening and nighttime hours tends to be associated with high annoyance, which is reflected in the use of community noise exposure metrics, such as the L_dn_ and L_den_, that include penalties during these hours. Transportation noise sources may exhibit distinct diurnal patterns, but the impact of these patterns on different noise metrics has not been thoroughly evaluated, especially within the United States. In this study, we utilized General Transit Feed Specification (GTFS) data from 24 major cities in the U.S. to quantify diurnal traffic patterns for local buses, and the impact of these patterns on differences in noise metrics, such as L_Day_, L_Evening_, L_Night_, L_dn_, and L_den_, compared to the 24-hour L_Aeq24_, Using mathematical conversions between the noise metrics, we found on average across the cities that the L_dn_ was between 2.8 to 3.6 dB higher than the L_Aeq24_, and the L_den_ was also 3.6 to 3.8 dB higher than the L_Aeq24_ for noise from local buses. This increase was mainly due to noise during daytime (L_Day_) that was higher than the 24-hour average noise, and dB penalties added to the L_dn_ and L_den_ metrics, which compensate for less bus traffic during evening and nighttime hours. We discuss the relevance of these conversions and the observed differences between the 24-hour L_Aeq24_ and the L_dn_ and L_den_, which are used for health impact assessments of high annoyance, on public transportation planning.

## Introduction

Transportation is an important contributor to noise in urban areas. An estimated 94.9 million people are exposed to transportation noise levels that exceed 45 dB in the United States (Seto & Huang, 2023).

Transportation noise is also an environmental justice concern because it disproportionately affects those who live along major transportation corridors. In the U.S., different population groups may be more impacted by certain types of transportation noise. One study found that Asian populations are most impacted by roadway traffic noise, Black populations most impacted by rail noise, and Hispanic populations most impacted by aviation noise (Huang & Seto, 2023; Seto & Huang, 2023). Transportation noise may vary considerably throughout the day, evening, and nighttime hours -- especially, roadway traffic-related noise, which may be driven by the hours and directional flow of commuter traffic. However, few studies have quantified the impact of diurnal traffic patterns on noise.

Various noise metrics have been developed to help characterize noise that exists during particular hours of the day. Compared to the L_Aeq24_ metric, which is the 24-hour average noise level, certain metrics, such as L_Day_, L_Evening_, and L_Night_ are used to quantify noise levels that are only experienced during certain hours of a 24-hour period. Furthermore, because noise during evening and nighttime hours may be more disturbing to people than noise during daytime hours, various metrics such as the day-night L_dn_ and day-evening-night L_den_ noise metrics were developed, which include penalties (5 dB during evening hours and 10 dB during nighttime hours). The L_dn_ and L_den_ are particularly relevant for population health and health impact assessments, as these metrics are typically used to estimate the percentage and numbers of persons in a population that may be highly annoyed by noise (Federal Interagency Committee on Noise, 1992; Schultz, 1978; World Health Organization, 2018). Conversions between these noise metrics is particularly relevant in the U.S. because the federal Bureau of Transportation Statistics has generated modeled national transportation noise levels, but only as L_Aeq24_ estimates (US Bureau of Transportation Statistics, 2022). Conversions from the L_Aeq24_ to other noise metrics such as the L_dn_ or L_den_ are generally needed in order to utilize existing exposure-response functions to estimate health impacts (Huang & Seto, 2023).

Previous work has derived mathematical relationships between various noise metrics (Brink et al., 2018). As Brink et al. explain, generally these relationships are not well understood or documented in either in the peer-reviewed scientific literature or policy and planning documents. The authors’ research examined empirical transportation data from the United Kingdom and Europe to quantify diurnal patterns and estimate dB differences between the various noise metrics. They found traffic noise tends to be elevated during daytime and evening hours, and observed L_dn_ and L_den_ metrics are higher than the L_Aeq24_ despite less traffic in nighttime hours. However, similar analyses have not been carried out in the U.S. Therefore, little evidence exists on how current diurnal traffic patterns relate to differences in noise metrics in the U.S.

Modern digital forms of traffic data may inform our understanding of diurnal traffic patterns. This is particularly the case for public transit traffic, which is a regular recurring source of noise along major transit corridors in urban areas. Transportation agencies have made their routes, schedules, stop locations, and trip stops available to the public in the form of General Transit Feed Specification (GTFS) files. The information contained in these files may be used to inform the relative proportions of public transit trips that occur during different hours of the day, and hence inform differences between various noise exposure metrics.

In this study, we utilized General Transit Feed Specification (GTFS) data from 24 major cities in the U.S. to quantify diurnal traffic patterns for local buses. Using the same noise metrics described by Brink, et al. (2018), we computed differences in noise metrics, such as L_Day_, L_Evening_, L_Night_, L_dn_, and L_den_, compared to the 24-hour L_Aeq24_. We hypothesized that similar to the findings for the UK and European countries, the penalized L_dn_ and L_den_ noise metrics will be higher than the L_Aeq24_ metric, even though local bus traffic may be considerably less during nighttime than daytime hours. We discuss the findings on the differences between noise exposure metrics for local bus traffic in terms of their impact on estimated high annoyance to noise in population health impact assessment studies in the U.S.

## Methods

### Noise Exposure Metrics

Below are the various noise exposure metrics considered in this study, as originally described by Brink et al. (2018). Notably, the L_Aeq24_ is the 24-hour average noise exposure, without any penalties added. L_Day_, L_Evening_, and L_Night_ are exposure metrics that account for noise exposure that occurs only during specific hours of the 24-hour period. L_dn_ and L_den_ exposure metrics are 24-hour averages that include 5 and 10 dB penalties during specific evening and nighttime hours, respectively. Also note that for the definition of some metrics there may be regional variations in the hours that are included for day, evening, versus night, which is reflected in the variations denoted with subscripts (e.g., a, b, c).

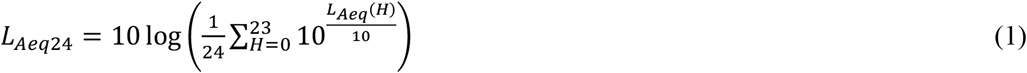

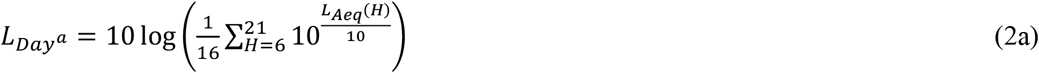

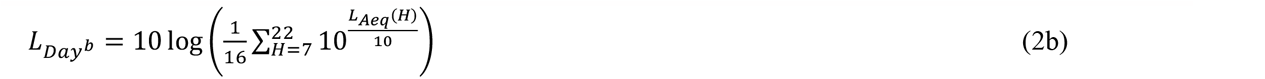

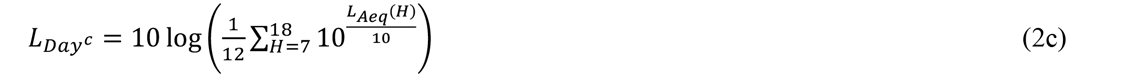

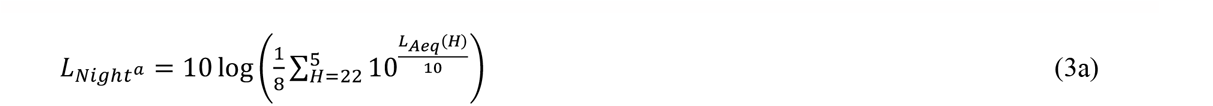

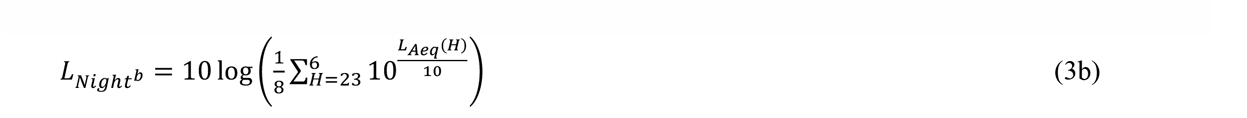

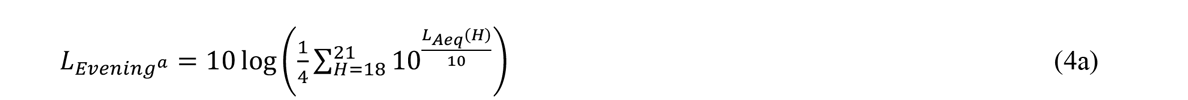

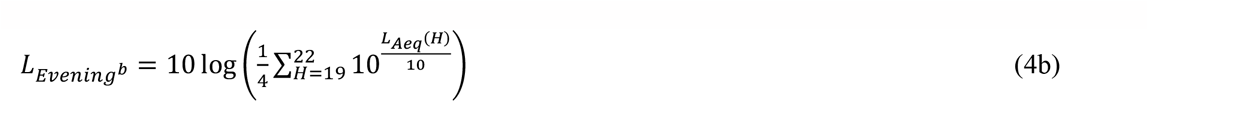

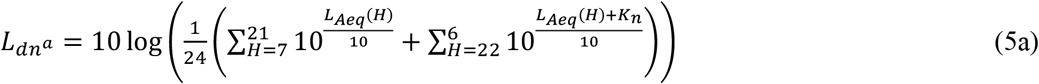

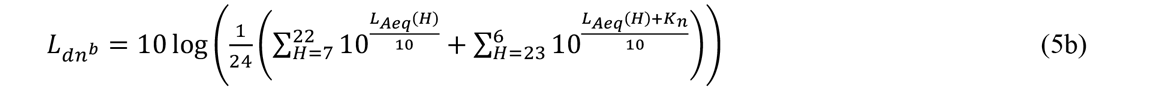

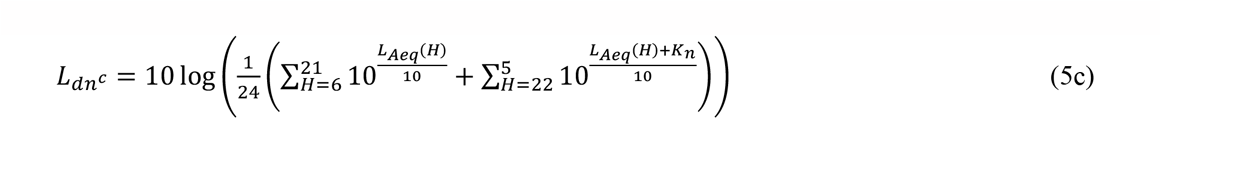

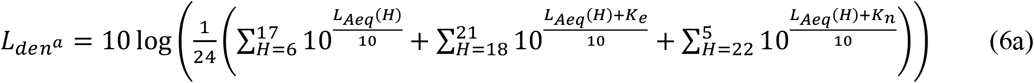

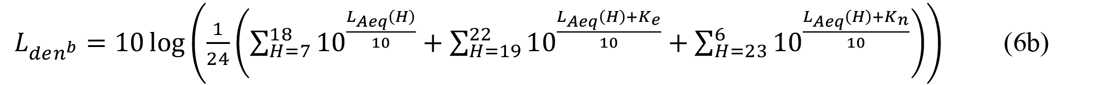

Above, *L*_*Aeg*_(*H*) represents the average noise level for the specific hour H. The penalties for the L_dn_ and L_den_ equations are represented by the K_n_ (K_n_=10 for the 10 dB nighttime penalty) and K_e_ (K_e_=5 for the 5 dB evening penalty).

### Deriving the Difference Between Noise Exposure Metrics

The difference between each noise metric and the 24-hour average (L_Aeq24_) can be derived algebraically as the difference in logs, or the log of the ratio. For example, the difference between the L_den_ and the L_Aeq24_ is shown below.

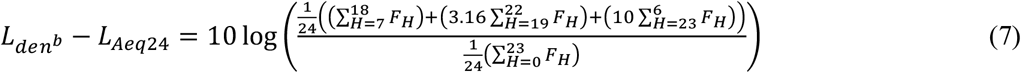

Above, F_H_ denotes the proportion of sound energy over the 24-hour period that occurs during hour H (i.e., 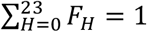. The factor of 3.16 represents that inclusion of the 5 dB penalty during the evening hours of H=[19,22] (i.e.,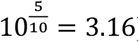) and the factor of 10 represents the 10 dB penalty during the nighttime hours of H=[23,6] (i.e.,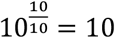) for the L_den_ metric. This relationship assumes “a bus is a bus” -- that sound energy per bus is similar across the hours of the day, which may not be the case for some bus routes and stops, as there may be differences in the proportion of certain types of buses or the speeds or other traffic/roadway factors could change with varying hours of the 24-hour period.

### Local Bus Traffic Data

GTFS data were obtained for 24 major cities in the U.S. to quantify diurnal traffic patterns for local buses. Generally, all GTFS datasets were available to the public and readily downloadable as compressed zip files from GTFS aggregator websites (MobilityData, 2023), with the exception of the San Francisco and Washington DC GTFS files, which required registering first for an Application Programming Interface (API), which subsequently allowed access to the transit agencies’ GTFS data (511 SF Bay, 2023; Washington Metropolitan Area Transit Authority, 2023). All GTFS data were downloaded on October 13, 2023.

For each city, we filtered the GTFS data to only include local bus schedules. Next, for each bus stop location, we computed the hourly number of bus visits to that location for each route schedule. We then summarized and visualized the diurnal distribution of bus visits across all the bus stop locations for each hour as a series of 24 box plots for each city.

To estimate the differences between each noise metric and the 24-hour average noise level (L_Aeq24_), we computed for each bus stop location, the proportion of bus visits over the 24-hour period that occur during each hour, which was used as a proxy for F_H_ in the noise difference equations (7). Average and standard deviations of the difference between each noise exposure metric and L_Aeq24_ were computed for each city using the difference equations. We also computed the average and standard deviations across the cities (i.e., mean of all the cities’ averages and SD of all the cities’ averages) to obtain a summary of the differences in noise metrics across the major cities of the U.S.

GTFS data were imported and processed in R (version 4.2.2) using the tidytransit (version 1.6.0) and dplyr (version 1.1.2) packages.

### Impact on High Annoyance in Population Health Impact Assessments

Because noise assessments in the U.S., such as the Bureau of Transportation Statistics’ national noise modeling work, may only produce estimates of L_Aeq24_, using the L_Aeq24_ instead of the L_dn_ or L_den_ in existing noise exposure-response functions may underestimate the percent population and numbers of population highly annoyed by transportation noise. To quantify the potential magnitude of this underestimate for noise from local buses, we applied the differences computed between the L_dn_ (and L_den_) and L_Aeq24_ to an existing noise exposure-response function (Miedema & Oudshoorn, 2001).

## Results and Discussion

The diurnal patterns of hourly bus visits to bus stops in each of the 24 U.S. cities is shown in Figure 1. Almost all of the cities demonstrate that bus traffic tends to be higher during the daytime hours, and is much lower during the nighttime hours, particularly around 2 a.m. However, we observe variations between cities, particularly in the magnitude of daytime bus visits per stop. Notably, large cities such as Chicago, New York, and San Francisco have higher numbers of bus visits per stop than other cities. This has potential relevance to bus traffic-induced noise exposures, as the greater amount of bus traffic in these cities (concentrated at particular bus stop locations), will contribute to a greater amount of noise-related health impacts.

**Figure 1.**
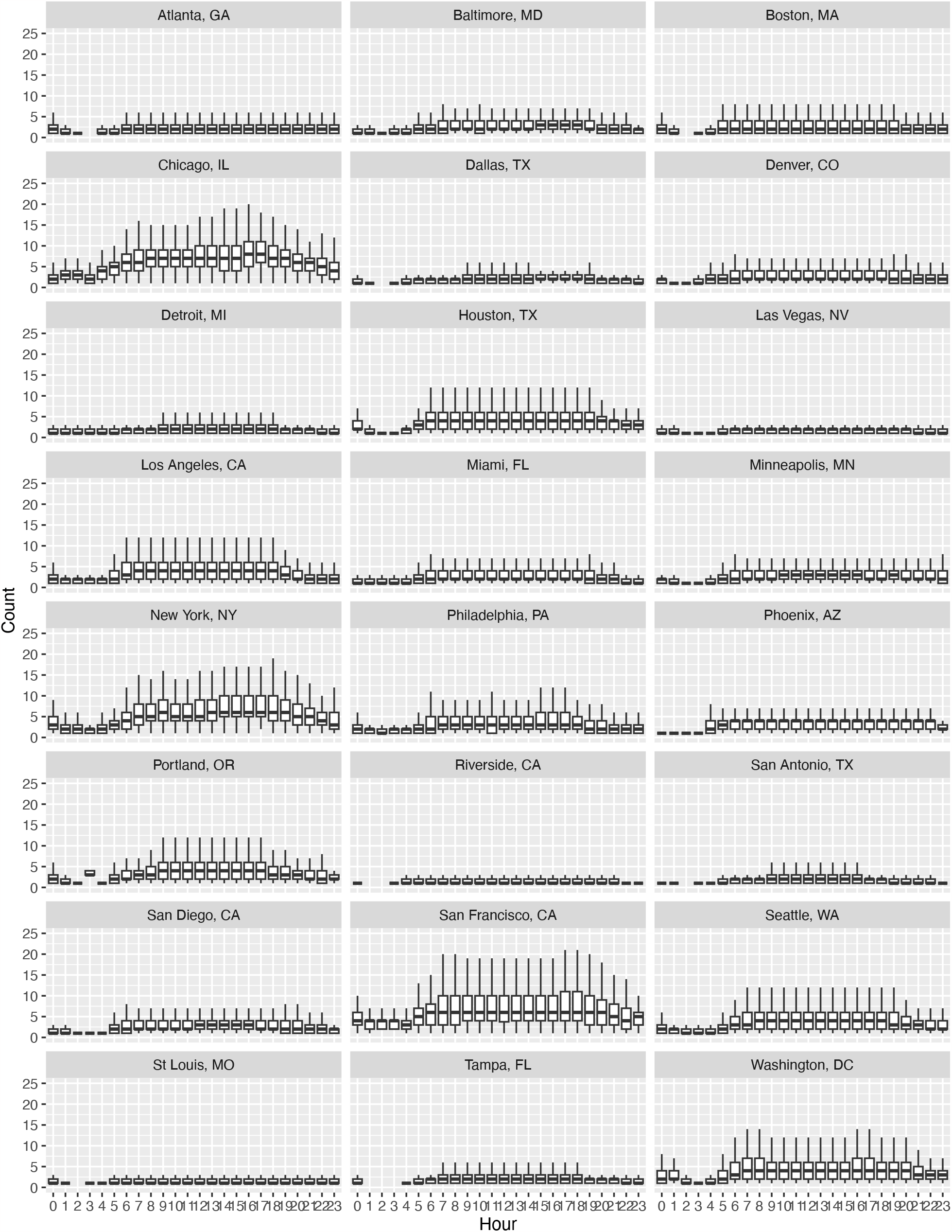
Boxplots of the diurnal pattern of the number local bus visits per bus stop in each city.

**Table 1** provides estimates of the differences (in dB) between the various noise metrics and the 24-hour average (L_Aeq24_). The standard deviations of the differences are provided in **Table 2**. We notice from **Table 1** that across all cities, the daytime noise metric (L_Day_) is higher (1.1 to 1.5 dB) than the L_Aeq24_. We also observe that the nighttime noise metric (L_Night_) is lower (3.7 dB) than the L_Aeq24_. Both of these findings are consistent with the diurnal traffic patterns illustrated in **Figure 1** that show higher daytime traffic than evening and nighttime traffic.

**Table 1.**
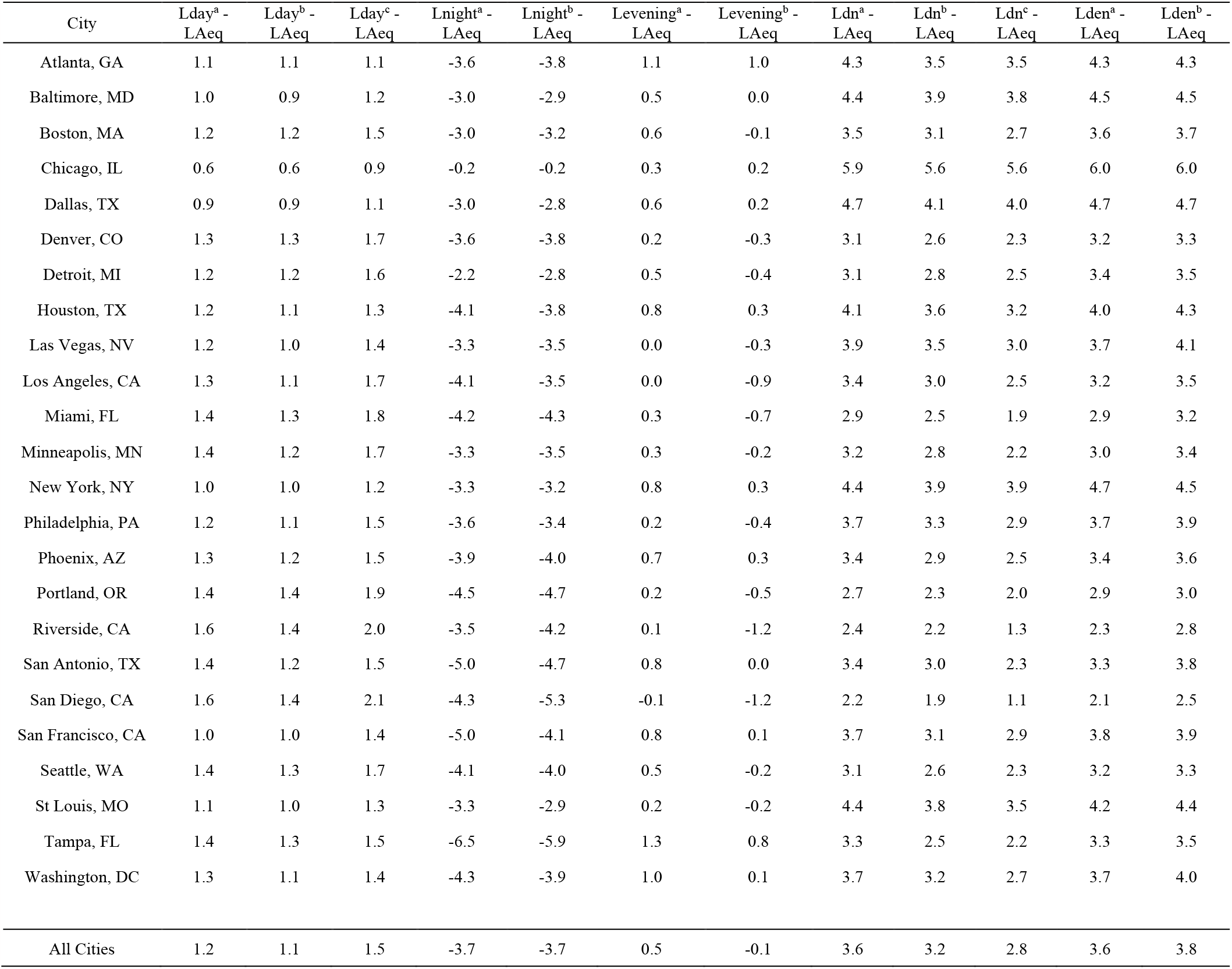
Average differences between 24-hour LAeq and other noise exposure metrics for local bus noise in US cities.

| City | Lday<sup>a</sup> - LAeq | Lday<sup>b</sup> - LAeq | Lday<sup>c</sup> - LAeq | Lnight<sup>a</sup> - LAeq | Lnight<sup>b</sup> - LAeq | Levening<sup>a</sup> - LAeq | Levening<sup>b</sup> - LAeq | Ldn<sup>a</sup> - LAeq | Ldn<sup>b</sup> - LAeq | Ldn<sup>c</sup> - LAeq | Lden<sup>a</sup> - LAeq | Lden<sup>b</sup> - LAeq |
| --- | --- | --- | --- | --- | --- | --- | --- | --- | --- | --- | --- | --- |
| Atlanta, GA | 1.1 | 1.1 | 1.1 | -3.6 | -3.8 | 1.1 | 1.0 | 4.3 | 3.5 | 3.5 | 4.3 | 4.3 |
| Baltimore, MD | 1.0 | 0.9 | 1.2 | -3.0 | -2.9 | 0.5 | 0.0 | 4.4 | 3.9 | 3.8 | 4.5 | 4.5 |
| Boston, MA | 1.2 | 1.2 | 1.5 | -3.0 | -3.2 | 0.6 | -0.1 | 3.5 | 3.1 | 2.7 | 3.6 | 3.7 |
| Chicago, IL | 0.6 | 0.6 | 0.9 | -0.2 | -0.2 | 0.3 | 0.2 | 5.9 | 5.6 | 5.6 | 6.0 | 6.0 |
| Dallas, TX | 0.9 | 0.9 | 1.1 | -3.0 | -2.8 | 0.6 | 0.2 | 4.7 | 4.1 | 4.0 | 4.7 | 4.7 |
| Denver, CO | 1.3 | 1.3 | 1.7 | -3.6 | -3.8 | 0.2 | -0.3 | 3.1 | 2.6 | 2.3 | 3.2 | 3.3 |
| Detroit, MI | 1.2 | 1.2 | 1.6 | -2.2 | -2.8 | 0.5 | -0.4 | 3.1 | 2.8 | 2.5 | 3.4 | 3.5 |
| Houston, TX | 1.2 | 1.1 | 1.3 | -4.1 | -3.8 | 0.8 | 0.3 | 4.1 | 3.6 | 3.2 | 4.0 | 4.3 |
| Las Vegas, NV | 1.2 | 1.0 | 1.4 | -3.3 | -3.5 | 0.0 | -0.3 | 3.9 | 3.5 | 3.0 | 3.7 | 4.1 |
| Los Angeles, CA | 1.3 | 1.1 | 1.7 | -4.1 | -3.5 | 0.0 | -0.9 | 3.4 | 3.0 | 2.5 | 3.2 | 3.5 |
| Miami, FL | 1.4 | 1.3 | 1.8 | -4.2 | -4.3 | 0.3 | -0.7 | 2.9 | 2.5 | 1.9 | 2.9 | 3.2 |
| Minneapolis, MN | 1.4 | 1.2 | 1.7 | -3.3 | -3.5 | 0.3 | -0.2 | 3.2 | 2.8 | 2.2 | 3.0 | 3.4 |
| New York, NY | 1.0 | 1.0 | 1.2 | -3.3 | -3.2 | 0.8 | 0.3 | 4.4 | 3.9 | 3.9 | 4.7 | 4.5 |
| Philadelphia, PA | 1.2 | 1.1 | 1.5 | -3.6 | -3.4 | 0.2 | -0.4 | 3.7 | 3.3 | 2.9 | 3.7 | 3.9 |
| Phoenix, AZ | 1.3 | 1.2 | 1.5 | -3.9 | -4.0 | 0.7 | 0.3 | 3.4 | 2.9 | 2.5 | 3.4 | 3.6 |
| Portland, OR | 1.4 | 1.4 | 1.9 | -4.5 | -4.7 | 0.2 | -0.5 | 2.7 | 2.3 | 2.0 | 2.9 | 3.0 |
| Riverside, CA | 1.6 | 1.4 | 2.0 | -3.5 | -4.2 | 0.1 | -1.2 | 2.4 | 2.2 | 1.3 | 2.3 | 2.8 |
| San Antonio, TX | 1.4 | 1.2 | 1.5 | -5.0 | -4.7 | 0.8 | 0.0 | 3.4 | 3.0 | 2.3 | 3.3 | 3.8 |
| San Diego, CA | 1.6 | 1.4 | 2.1 | -4.3 | -5.3 | -0.1 | -1.2 | 2.2 | 1.9 | 1.1 | 2.1 | 2.5 |
| San Francisco, CA | 1.0 | 1.0 | 1.4 | -5.0 | -4.1 | 0.8 | 0.1 | 3.7 | 3.1 | 2.9 | 3.8 | 3.9 |
| Seattle, WA | 1.4 | 1.3 | 1.7 | -4.1 | -4.0 | 0.5 | -0.2 | 3.1 | 2.6 | 2.3 | 3.2 | 3.3 |
| St Louis, MO | 1.1 | 1.0 | 1.3 | -3.3 | -2.9 | 0.2 | -0.2 | 4.4 | 3.8 | 3.5 | 4.2 | 4.4 |
| Tampa, FL | 1.4 | 1.3 | 1.5 | -6.5 | -5.9 | 1.3 | 0.8 | 3.3 | 2.5 | 2.2 | 3.3 | 3.5 |
| Washington, DC | 1.3 | 1.1 | 1.4 | -4.3 | -3.9 | 1.0 | 0.1 | 3.7 | 3.2 | 2.7 | 3.7 | 4.0 |
| All Cities | 1.2 | 1.1 | 1.5 | -3.7 | -3.7 | 0.5 | -0.1 | 3.6 | 3.2 | 2.8 | 3.6 | 3.8 |

**Table 2.**
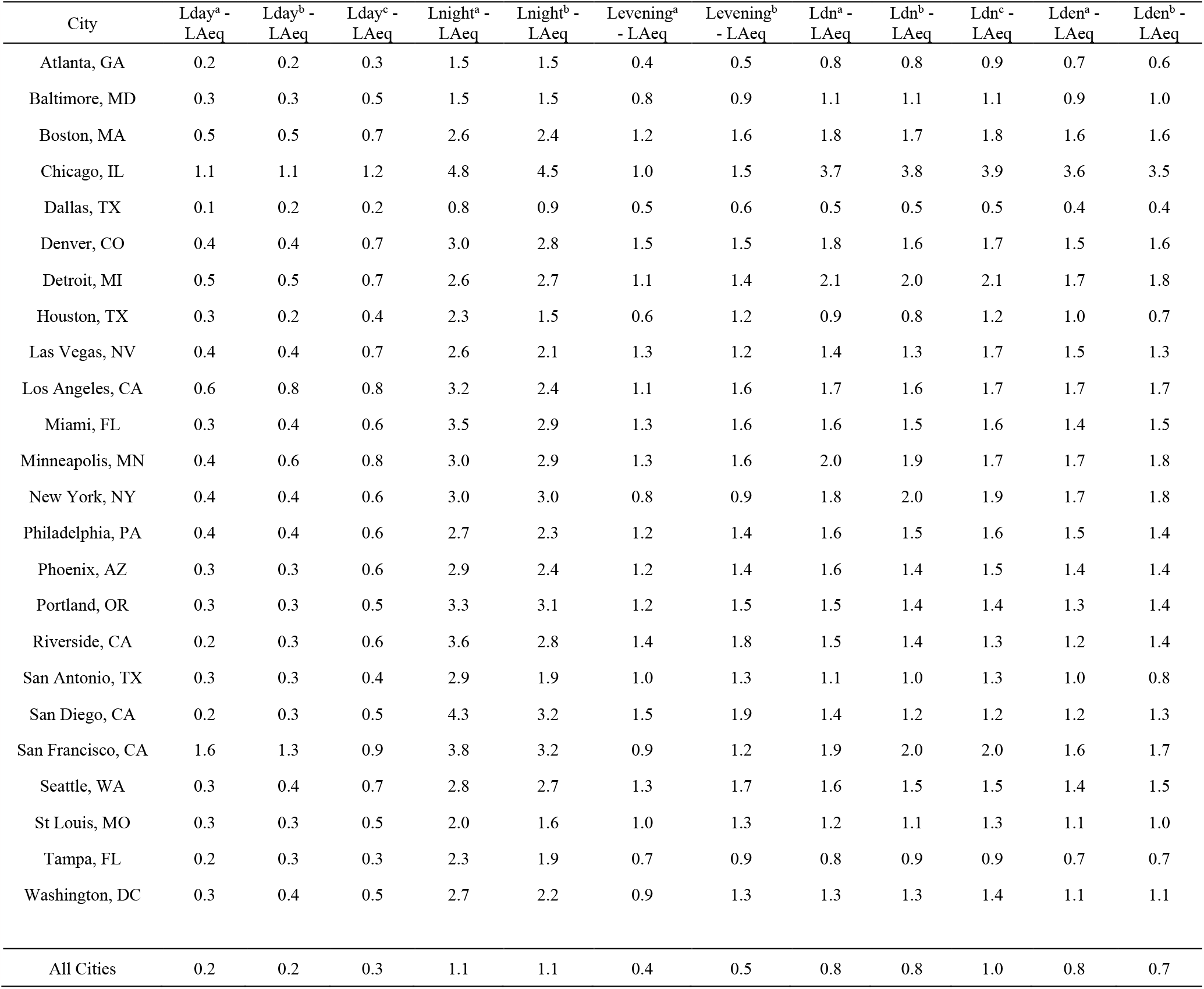
Standard deviations of the differences between 24-hour LAeq and other noise exposure metrics for local bus noise in US cities.

Interestingly, the evening metric (L_Evening_) can be either higher by on average 0.5 dB or lower by on average 0.1 dB than the L_Aeq24_ depending on which definition of L_Evening_ is used. The difference in definition (equations 4a and 4b) is a 1-hour shift to the later hours in L_Evening_^b^ compared to L_Evening_^a^. The shift to the later hours, corresponds to less bus traffic, which in turn, corresponds to a L_Evening_ that is slightly lower than the L_Aeq24_ when averaged across the U.S. cities.

For the penalized noise metrics, both the L_dn_ and L_den_ are higher than the L_Aeq24_. This is the case across all the cities we analyzed. Depending on the definition, and which hours are included as nighttime, the L_dn_ ranges on average across the cities, 2.8 to 3.6 dB higher than the L_Aeq24_. The L_den_ ranges on average across the cities, 3.6 to 3.8 dB higher than the L_Aeq24_. This confirms our hypothesis that the diurnal patterns of local bus-related noise in major U.S. cities results in L_dn_ and L_den_ metrics that are higher than the 24-hour average noise level (L_Aeq24_).

To gain better insights into why the L_dn_ and L_den_ metrics are higher than the L_Aeq24_, especially when the L_Day_ tends to be higher than the L_Aeq24_ and the L_Night_ tends to be lower than the L_Aeq24_, we computed some additional differences measures (**Table 3** shows average differences in dB, while **Table 4** shows the standard deviations of the differences). These difference measures include the 5 dB penalties for evening hours, and the 10 dB penalties for nighttime hours in the calculation of L_Evening_ and L_Night_, respectively.

**Table 3.** Average differences between 24-hour LAeq and Levening and Lnight with 5 and 10 dB penalties, <u>respectively, for local bus noise in US cities.</u>.

| City | Lnight<sup>a</sup> - LAeq | Lnight<sup>b</sup> - LAeq | Levening<sup>a</sup> - LAeq | Levening<sup>b</sup> - LAeq |
| --- | --- | --- | --- | --- |
| Atlanta, GA | 6.1 | 6.1 | 6.0 | 5.9 |
| Baltimore, MD | 6.6 | 6.8 | 5.4 | 4.8 |
| Boston, MA | 4.6 | 5.2 | 5.4 | 4.4 |
| Chicago, IL | 8.5 | 8.5 | 2.9 | 2.8 |
| Dallas, TX | 7.0 | 7.1 | 5.6 | 5.2 |
| Denver, CO | 3.7 | 4.4 | 5.0 | 4.1 |
| Detroit, MI | 4.2 | 4.7 | 5.4 | 4.3 |
| Houston, TX | 5.3 | 6.2 | 5.8 | 5.2 |
| Las Vegas, NV | 5.0 | 6.0 | 4.9 | 4.2 |
| Los Angeles, CA | 4.0 | 5.1 | 4.4 | 3.5 |
| Miami, FL | 2.9 | 4.1 | 5.1 | 3.9 |
| Minneapolis, MN | 3.6 | 4.6 | 4.7 | 3.7 |
| New York, NY | 6.5 | 6.4 | 5.4 | 4.9 |
| Philadelphia, PA | 4.8 | 5.5 | 5.1 | 4.2 |
| Phoenix, AZ | 4.1 | 4.8 | 5.4 | 4.8 |
| Portland, OR | 3.0 | 3.6 | 4.9 | 3.9 |
| Riverside, CA | 1.9 | 3.5 | 5.0 | 3.2 |
| San Antonio, TX | 3.6 | 5.1 | 5.8 | 4.9 |
| San Diego, CA | 1.3 | 2.9 | 4.6 | 3.0 |
| San Francisco, CA | 4.3 | 5.1 | 5.6 | 4.9 |
| Seattle, WA | 3.8 | 4.3 | 5.1 | 4.1 |
| St Louis, MO | 6.0 | 6.7 | 5.2 | 4.7 |
| Tampa, FL | 3.2 | 4.0 | 6.2 | 5.8 |
| Washington, DC | 4.5 | 5.5 | 5.9 | 5.0 |
| All Cities | 4.5 | 5.3 | 5.2 | 4.4 |

**Table 4.** Standard deviations of the differences between 24-hour LAeq and Levening and Lnight with 5 and 10 dB penalties, respectively, for local bus noise in US cities.

| City | Lnight<sup>a</sup> - LAeq | Lnight<sup>b</sup> - LAeq | Levening<sup>a</sup> - LAeq | Levening<sup>b</sup> - LAeq |
| --- | --- | --- | --- | --- |
| Atlanta, GA | 1.7 | 1.6 | 0.5 | 0.7 |
| Baltimore, MD | 1.9 | 1.9 | 1.0 | 1.2 |
| Boston, MA | 3.2 | 3.0 | 1.6 | 2.2 |
| Chicago, IL | 5.8 | 5.6 | 3.0 | 3.1 |
| Dallas, TX | 0.9 | 1.0 | 0.5 | 0.7 |
| Denver, CO | 3.1 | 3.0 | 1.8 | 2.2 |
| Detroit, MI | 3.7 | 3.5 | 1.3 | 1.8 |
| Houston, TX | 2.4 | 1.5 | 0.8 | 1.3 |
| Las Vegas, NV | 3.1 | 2.4 | 1.5 | 1.9 |
| Los Angeles, CA | 3.1 | 2.8 | 2.0 | 2.1 |
| Miami, FL | 3.0 | 2.8 | 1.7 | 2.0 |
| Minneapolis, MN | 3.2 | 3.3 | 2.3 | 2.5 |
| New York, NY | 3.1 | 3.2 | 1.7 | 1.7 |
| Philadelphia, PA | 3.0 | 2.8 | 1.5 | 1.8 |
| Phoenix, AZ | 3.0 | 2.6 | 1.8 | 2.1 |
| Portland, OR | 2.7 | 2.8 | 1.7 | 2.1 |
| Riverside, CA | 2.3 | 2.5 | 1.6 | 2.1 |
| San Antonio, TX | 2.5 | 2.0 | 1.1 | 1.4 |
| San Diego, CA | 2.3 | 2.4 | 1.9 | 2.2 |
| San Francisco, CA | 3.8 | 3.3 | 1.5 | 1.6 |
| Seattle, WA | 2.7 | 2.9 | 1.9 | 2.3 |
| St Louis, MO | 2.4 | 2.0 | 1.1 | 1.6 |
| Tampa, FL | 2.1 | 1.9 | 0.8 | 1.0 |
| Washington, DC | 2.7 | 2.4 | 1.2 | 1.6 |
| All Cities | 1.7 | 1.3 | 0.7 | 0.8 |

With the penalties included, we can see that both the penalized L_Evening_ and L_Night_ metrics are both higher than the L_Aeq24_, and therefore contribute to why both the L_dn_ and L_den_ metrics are higher than the L_Aeq24._ Even though bus traffic is lower during the nighttime, the 10 dB penalty (as well as the 5 dB penalty for evening hours) results in higher L_dn_ and L_den_ than the L_Aeq24_.

Do these differences matter in terms of health impacts? In the U.S., because the national noise models estimate L_Aeq24_, we might be tempted to use the L_Aeq24_ as a surrogate for either the L_dn_ or L_den_ in noise health impact assessment calculations. However, we can see from our results (**Table 1**) that doing so might underestimate the L_dn_ or L_den_ by between 2.8 to 3.8 dB. **Figure 2** shows the noise exposure-response curve for percent high annoyance to roadway transportation noise, with shifts shown in shaded areas under the assumption of L_dn_ = L_Aeq24_ versus the assumption of L_dn_ = (L_Aeq24_ + 3 dB). Due to the non-linear curve in the exposure-response function, the impact of the 3 dB difference on the percent highly annoyed ranges from 1.4 % at relatively low traffic noise levels (1.5 % HA for 45 dB vs 2.9 % HA for 48 dB) to 6.8 % at relatively high traffic noise levels (25.1 % HA at 70 dB vs 31.8 % HA at 73 dB).

**Figure 2.**
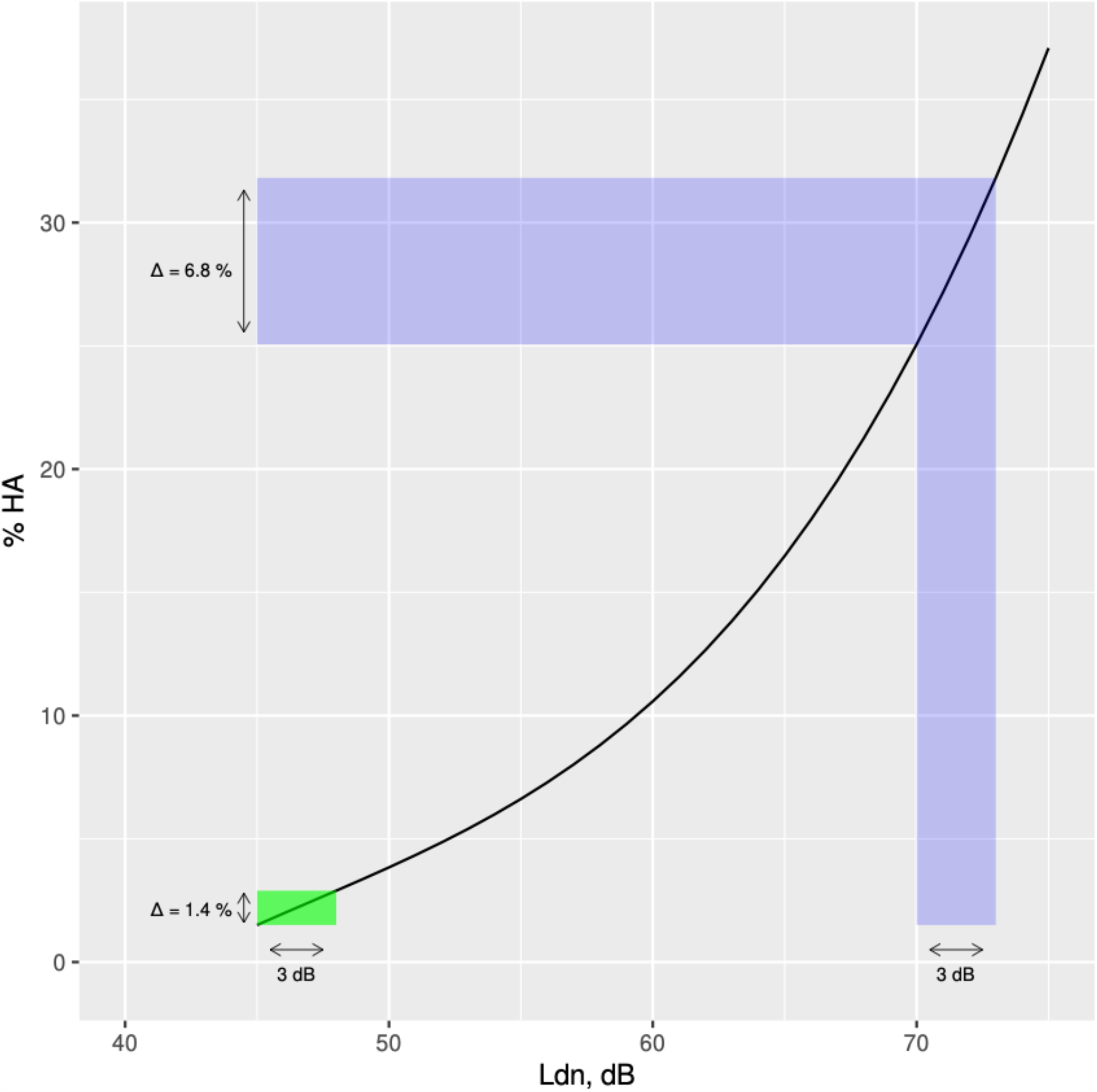
Road traffic exposure-response curve with differences in the percent highly annoyed shown for 3 dB differences in Ldn: in green for 45-48 dB difference, and in blue for 70-73 dB difference.

Multiplied by the large population numbers across all urban communities in the U.S., even these small differences in the percent highly annoyed result in large differences in the numbers of people highly annoyed by local bus noise. The larger percentage differences at higher transportation noise levels (e.g., the 6.8% difference at 70-73 dB) may be particularly problematic for cities such as Chicago, New York, and San Francisco that have relatively large numbers of bus visits per bus stop location.

## Conclusions

This study is perhaps one of the few analyses considering diurnal traffic patterns for buses in U.S. cities, and the impact of these diurnal patterns on various noise exposure metrics compared to the 24-hour average noise level (L_Aeq24_). The analyses focused on hourly local bus traffic data from GTFS files. GTFS data, while not new, are not often used in noise-related analyses, and thus, are a useful addition to the sources of traffic data that can be used to characterize and quantify traffic magnitudes and variations between cities, at different transit stop locations, and for different times of the day. By computing the differences between noise metrics, we found that the penalized noise metrics (L_dn_ and L_den_), which are often used in health impact assessments are higher than the L_Aeq24_ for local bus diurnal traffic patterns.

The approximate 3 dB difference penalized noise metrics versus the L_Aeq24_ should be considered when computing health impacts in transportation-related noise assessments, particularly if using exposure-response curves that exhibit non-linear larger effects per dB increment at higher noise levels, and when conducting assessment for cities with those higher noise levels. Furthermore, our findings suggest that local city buses to have the potential to contribute to transportation-induced high annoyance, as the higher L_dn_ and L_den_ compared to the L_Aeq24_ indicate that the dB penalties more than compensates for the relatively less bus traffic during nighttime hours in U.S. cities.

## Data Availability

All data produced in the present study are available upon reasonable request to the authors

